# Barriers and facilitators to specialist service referrals for sleep disorders from healthcare professionals’ perspective: A scoping review of qualitative research evidence

**DOI:** 10.1101/2025.01.20.25320835

**Authors:** Martina Sýkorová, Ellen Nolte, Frederick Martineau, Michelle A. Miller, Sofia H. Eriksson, Ian E. Smith, Helen Strongman

## Abstract

**Background:** Access to specialist sleep centres can lead to timely diagnosis and management of sleep disorders, improving the health and quality of life of people with these conditions. Evidence describing factors that influence referral to specialist sleep services is needed to inform improvements to the referral process.

**Objective:** This scoping review identifies and summarises qualitative evidence on barriers and facilitators to referrals to specialist sleep services by healthcare professionals.

**Eligibility criteria:** The review considered qualitative studies published in any language between 1995-2024 that explore the barriers and/or facilitators to referral to specialist services for sleep-related disorders from healthcare care professionals’ perspective.

**Sources of evidence:** A systematic search of six databases (Cochrane Library, MEDLINE, PubMed, Scopus, EMBASE and Open Grey) was conducted.

**Results:** Out of 3507 records, seven heterogeneous studies from two countries (Australia and USA) and one multi-country study met the inclusion criteria. Referrals were from a range of settings including primary care (two studies). Major themes identified as influencing referral included clarity of roles, referral experiences and options, knowledge of sleep disorders, funding, and views on treatment. Specific barriers and facilitators varied between countries, healthcare professionals and sleep disorders.

**Conclusions:** Although specific barriers and facilitators vary between settings, cross-cutting themes influencing referral to sleep centres reflect a lack of awareness of sleep disorders among healthcare professionals, poorly defined referral pathways, and funding issues.

**Impacts:** This is the first scoping review to synthesise qualitative evidence on factors influencing referrals to specialist services for sleep disorders. The themes identified can be used in further research to inform improvements to referral pathways in different settings.

**Reporting Method:** Adherence to the PRISMA-ScR checklist was maintained.

## 1. Introduction

Sleep disorders are a heterogenous group of conditions of diverse aetiology. Although sleep disorders are increasingly recognised as a public health problem worldwide, they remain under-recognised, under-diagnosed and under-treated (1). Early diagnosis and treatment by a sleep specialist can reduce many of the negative impacts including poor educational outcomes, unintentional injuries such as road traffic and other accidents as well as wider productivity losses in the workplace (1,2).

In many countries, people needing access to specialist services for conditions such as sleep disorders require a referral from another healthcare professional (3–6), in most cases their general practitioner (GP) (5,7–9). Some studies suggest that this form of primary care gatekeeping may be associated with delayed diagnosis and, ultimately, poorer outcomes for conditions such as cancer (10) and rare diseases (11). Challenges that have been cited include: interpretation of symptoms and readiness to further investigate (12,13); lack of confidence among primary care practitioners making referral decisions and limited understanding of and links to specialist care providers, along with time pressures and workload (9). Little is known about the factors that influence referrals to specialist sleep services; this information would improve the diagnosis process and access to appropriate care.

We conducted a preliminary search of MEDLINE, the Cochrane Database of Systematic Reviews and *JBI Evidence Synthesis* to confirm that there are no current or underway systematic or scoping reviews exploring factors influencing referrals to specialist sleep services. This paper therefore presents a scoping review that summarises the qualitative evidence on factors that influence referrals to specialist services for sleep-related disorders. We focus on the perspective of healthcare providers.

## 2. Methods

We used the Participants/Concept/Context framework, as recommended by the Joanna Briggs Institute for scoping reviews (14), to structure the primary objective and define the inclusion criteria.

**Figure 1:**
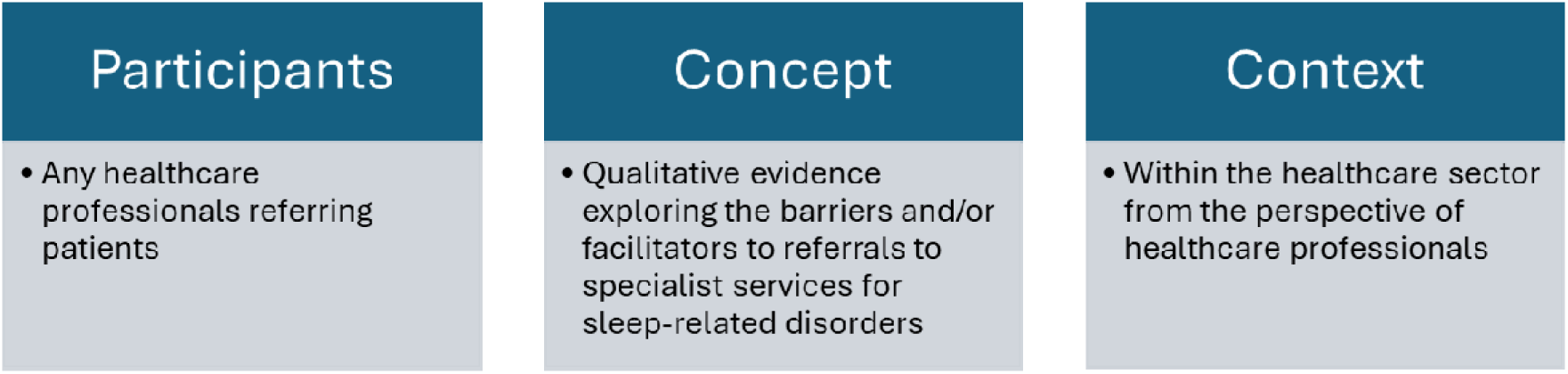
Research question generation.

The proposed scoping review was conducted in accordance with the JBI methodology for scoping reviews (15) and reported using the PRISMA-ScR Checklist (see appendix A) (16).

### 2.1. Protocol and Registration

A protocol was developed and is included in appendix B. We did not register the protocol as PROSPERO, an international database of prospectively registered reviews, does not accept scoping review protocols for registration (17).

### 2.2. Eligibility criteria

Table 1 summarises the eligibility criteria. We included studies published since 1995 to capture major developments in sleep medicine (18) and our understanding of sleep disorders, namely (i) the introduction of commercial continuous positive airway pressure (CPAP) machines in 1995 for the treatment of sleep apnoea (19), (ii) the identification of hypocretin deficiency as the cause of narcolepsy in 2000 (20), and (iii) discoveries in the treatment of insomnia in the 1990s (21). As this scoping review focuses on the multi-faceted and complex factors influencing referrals, as opposed to pre-identified barriers or enablers, all types of qualitative studies were included.

**Table 1:** Eligibility criteria.

| Inclusion criteria | Exclusion criteria |
| --- | --- |
| **View:** Referrals from any healthcare professional | **View:** Patients', family members' or carers' perspective |
| **Types of studies:**<br>Any qualitative methodologies (including qualitative studies as part of mixed method studies)<br>Systematic reviews | **Types of studies:**<br>Quantitative methodologies<br>Reviews, expert opinions, abstracts, editorials, comments, study protocols or studies with no full text available |
| **Language:** Any |  |
| **Outcome:** Reporting on barriers and/or facilitators to referrals for sleep-related disorders |  |
| **Referral to:** specialist sleep/ respiratory/ neurology/ psychiatry services for sleep-related disorders |  |
| **Years:** 1995-2024 |  |

### 2.3. Search strategy

We conducted a phased search strategy. We first carried out an initial search using two databases MEDLINE (PubMed) and Google Scholar to identify relevant articles and used words contained in the titles and abstracts of identified articles and the index terms used to describe these articles to develop a full search strategy. We consulted with a librarian to advise on the proposed search strategy (for the full search strategy please see appendix C). In a second step, we adapted the search strategy, including all identified keywords and index terms, to each included database and/or information source. We searched the following databases: Cochrane Library, MEDLINE (via Ovid), PubMed, EMBASE (via Ovid), Scopus and Open Grey for sources of grey literature. Finally, we screened the reference lists of all included sources of evidence for additional eligible studies. We included one additional study that was identified outside the formal search when reviewing literature for the discussion section of this review. This manuscript was published in 2024 and is not yet indexed in any of the included databases. No automation tools were used to remove any citations.

### 2.4. Selection of sources of evidence

One reviewer screened all the abstracts and full texts (MS). For studies where eligibility was uncertain, additional reviewers (HS, FM) were consulted and a mutual decision agreed as to their eligibility. We used the Rayyan systematic review platform to manage the review process (22). Screening was done after duplicates were removed.

### 2.5. Data extraction

We piloted and revised the extraction tool using five studies before beginning formal screening. Data were extracted from included papers by one reviewer (MS) using the data extraction tool mentioned above (appendix D). Extracted data included: study characteristics (authors, country of origin, year of publication), type of study (methodology, sample size, participant information), and barriers and/or facilitators to referrals to specialist sleep services.

### 2.6. Data synthesis

We grouped studies by the types of barriers and/or facilitators identified by study authors, and summarised the type of healthcare settings, sleep disorders and referring healthcare providers, along with the qualitative findings. To make sense of the qualitative data, we used thematic analysis to identify, analyse, and interpret themes (23). The excerpts of text of included studies were examined and coded. Initial codes were developed iteratively through team discussion. Grouping of the refined codes allowed for the emergence of patterns and overarching themes that helped us to address the research objective.

## 3. Results

### 3.1. Selection of sources of evidence

In total, the database searches identified 3507 records. After duplicates were removed (n = 488), 3019 abstracts were screened, of which fourteen were eligible for a full-text review. Six studies met the inclusion criteria. A hand search of reference lists of included studies did not identify any additional studies. However, a further literature search for the purpose of writing the discussion part of this paper identified an additional study that met the inclusion criteria. In total, seven studies were included for narrative review as summarised in figure 2.

**Figure 2:**
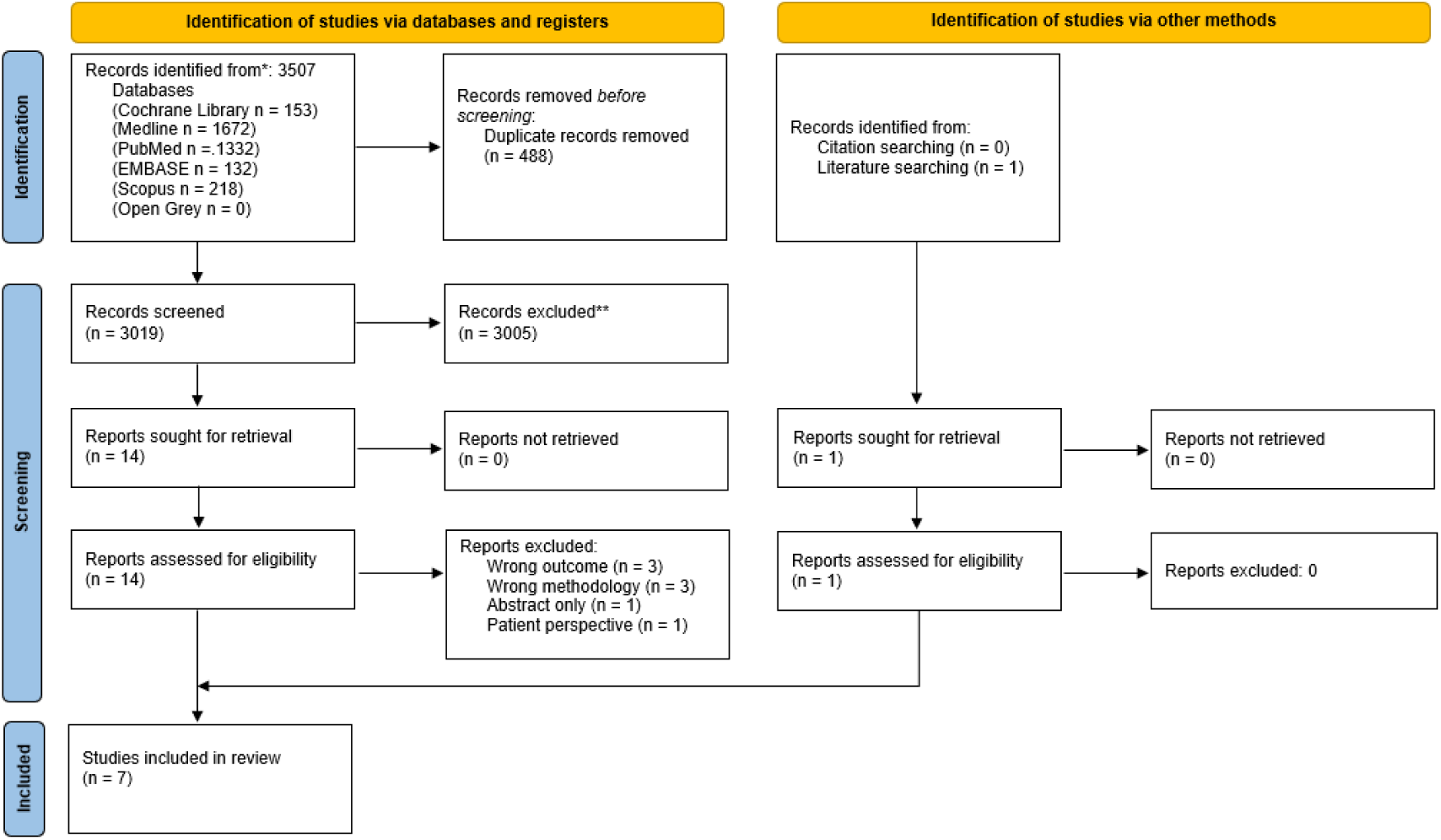
PRISMA diagram.

### 3.2. Characteristics of sources of evidence

Out of the included seven studies, one study was published in 2012 (24) and the remaining six studies were published in the past six years (2019–2024) (25–30). Three of the included studies were set in the USA (24–26), three in Australia (28–30), and one study looked at healthcare staff perspectives in the UK, Canada, USA, Australia and New Zealand (27). Table 2 summarises the included studies and main findings.

**Table 2:**
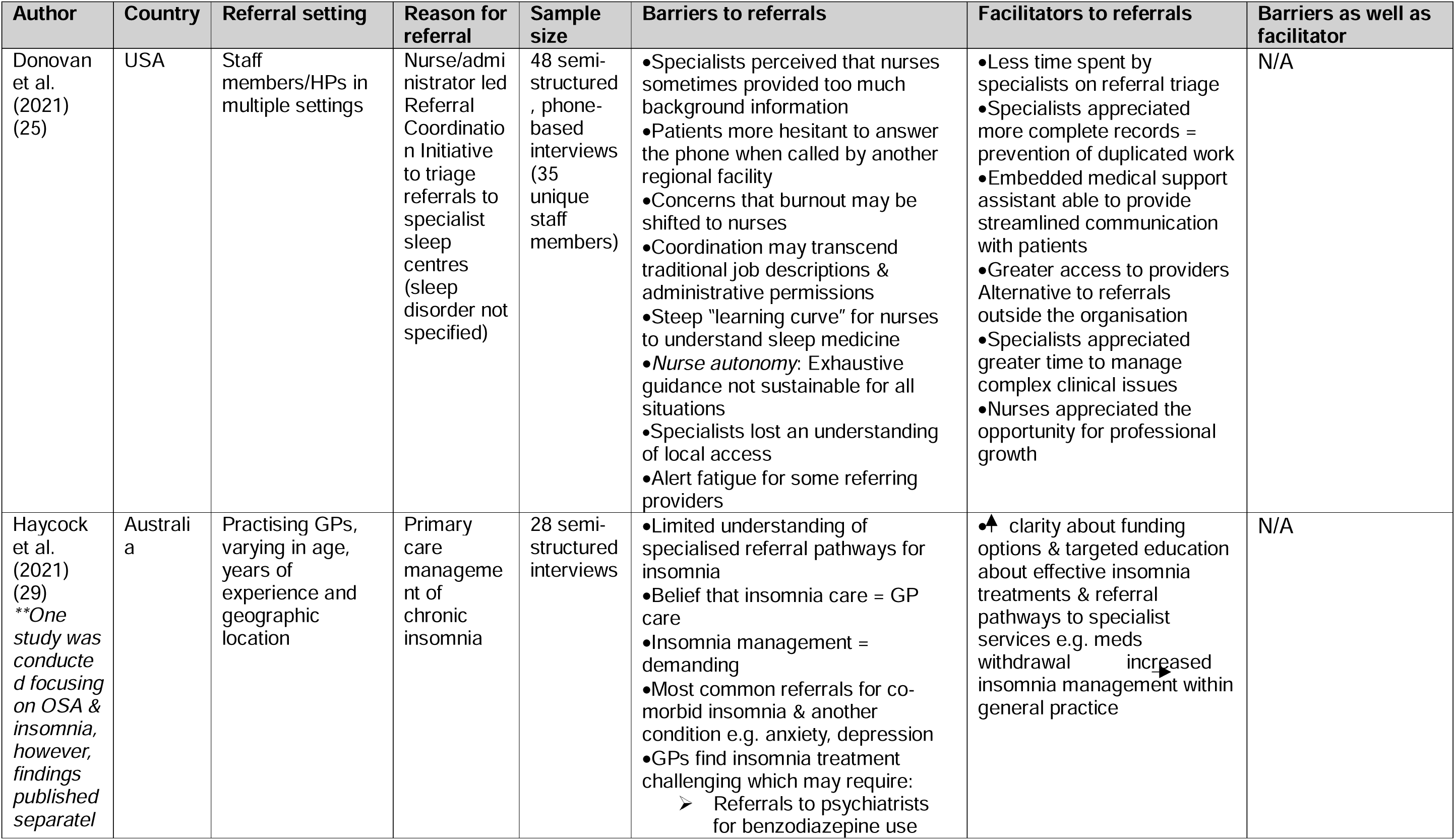
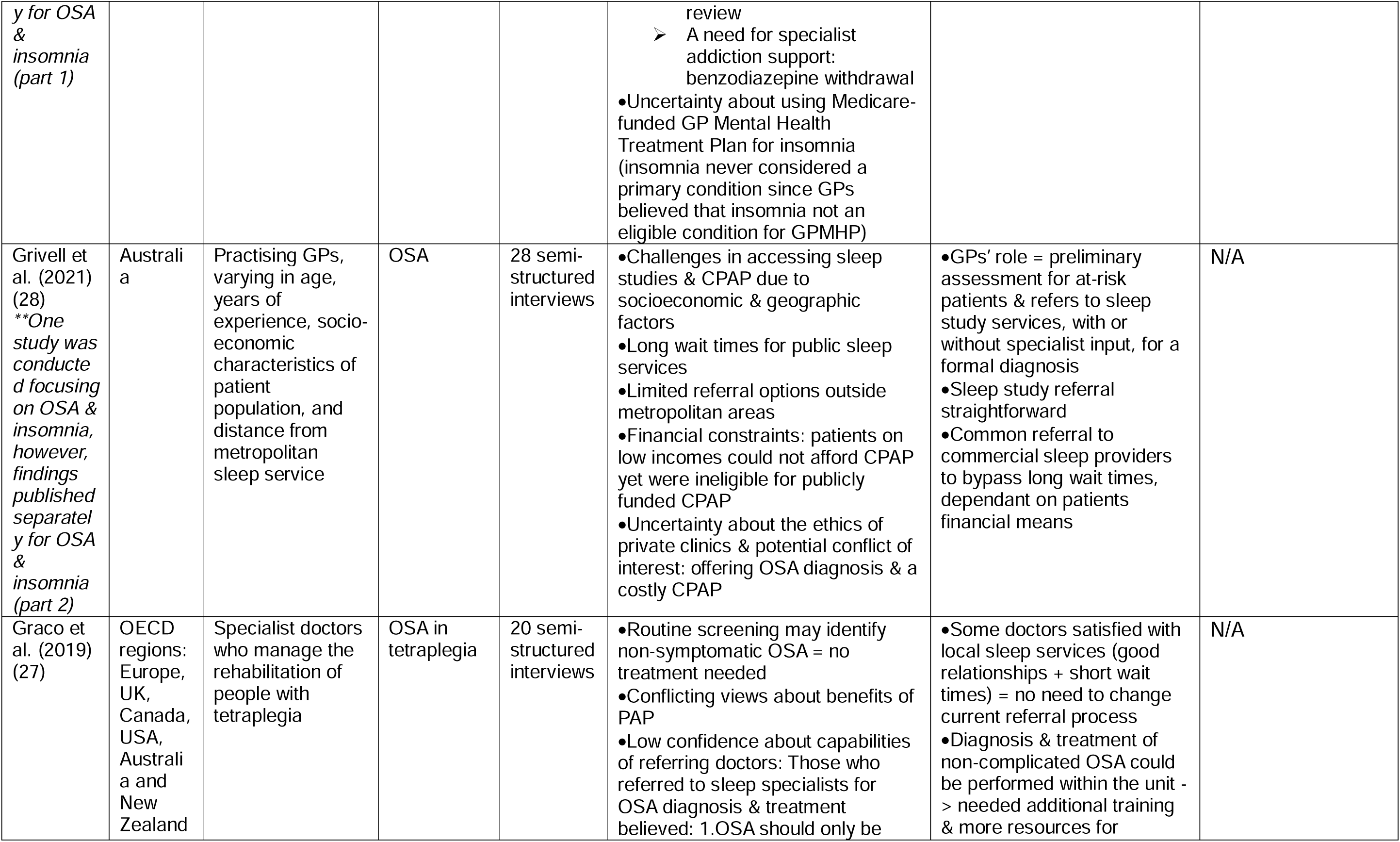
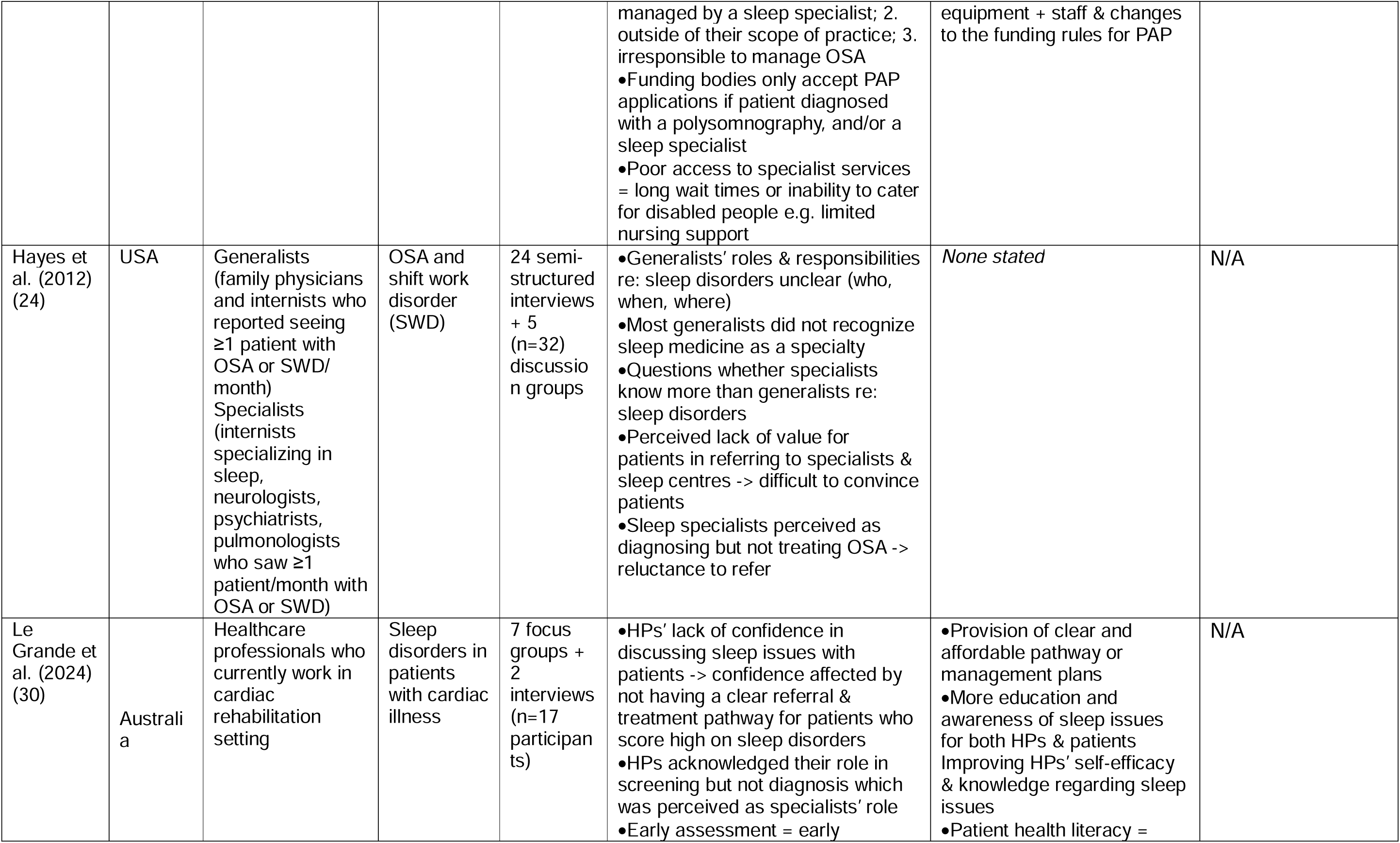
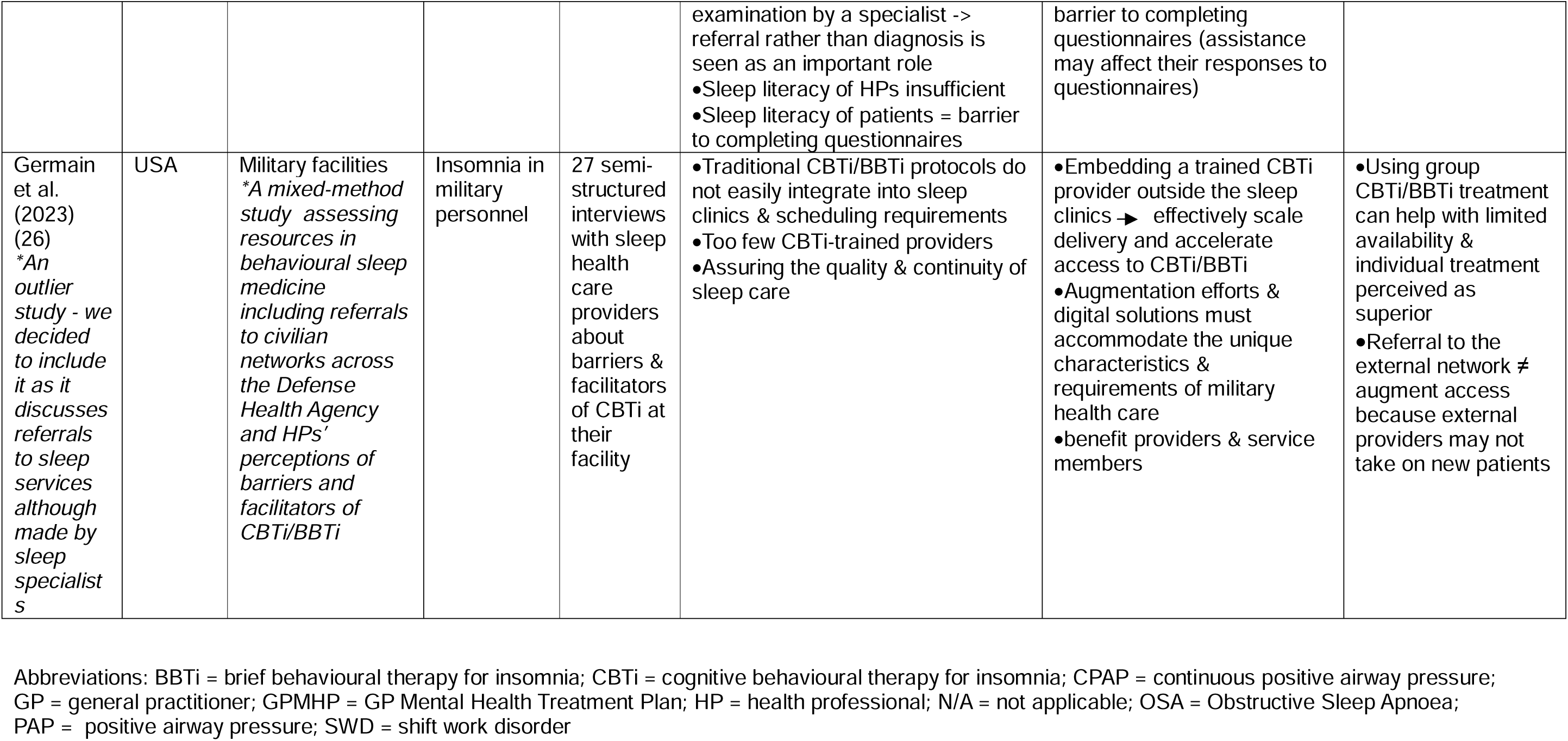
Included studies and findings.

Included studies explored views on referral from the perspectives of: GPs (28,29), staff working in military facilities (26), specialist doctors managing people with tetraplegia (27), healthcare professionals practising in cardiac rehabilitation (30), generalists (family physicians and internists) as well as specialists (24), and multiple staff (25). Two studies focused on referral for suspected symptoms of OSA (27,28), one study explored OSA and shift work disorder (24), two studies investigated insomnia (26,29), and two studies focused on unspecified sleep disorders including OSA and insomnia (25,30). All included studies utilised semi-structured interviews (n= 2-48), two studies in conjunction with focus group discussions with two to six participants per group (24,30). Table 3 provides a summary of the barriers and facilitators identified in this scoping review.

**Table 3:**
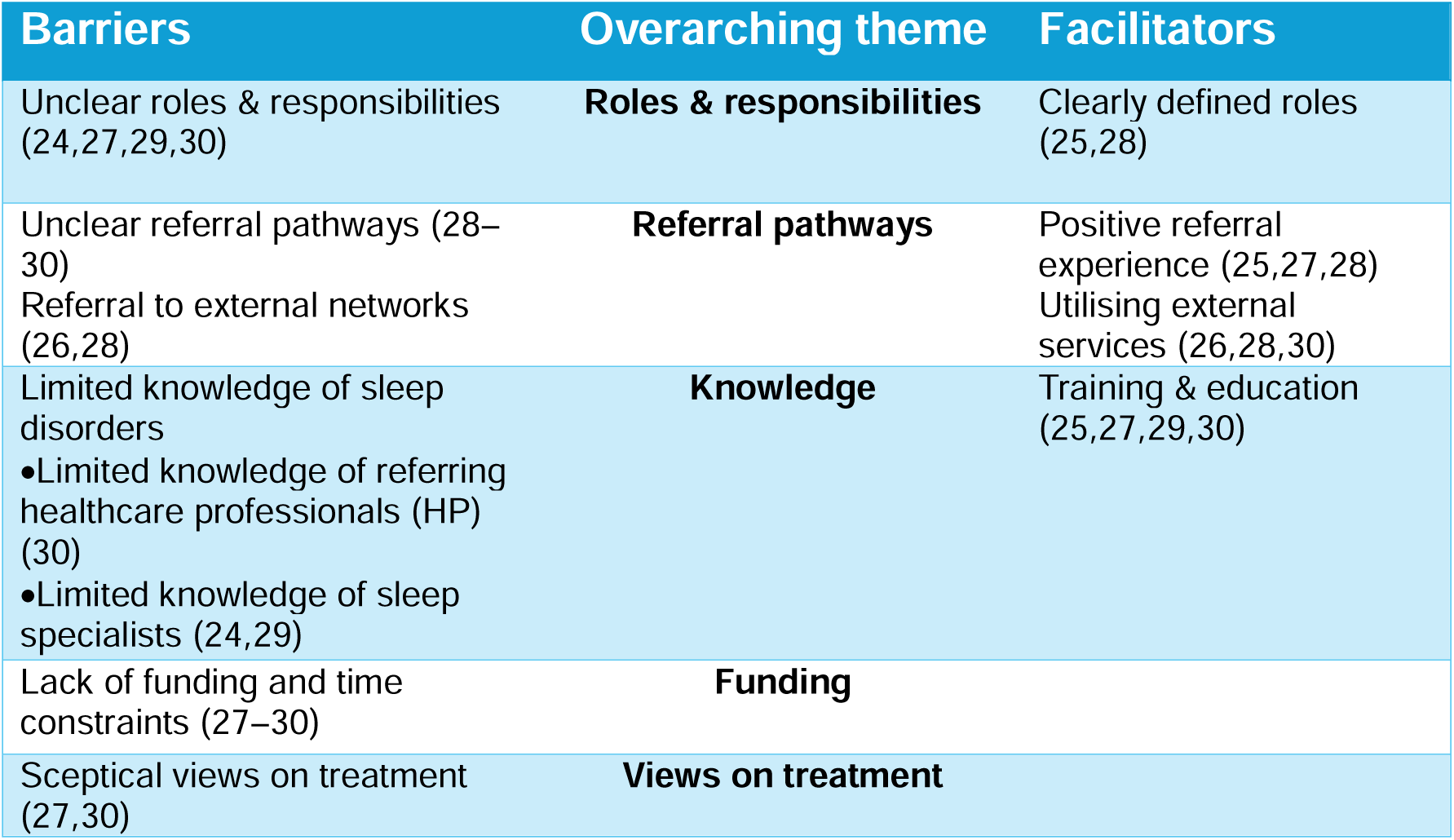
Summary of themes.

### 3.3. Overview of findings

We identified the following barriers to sleep specialist referrals: unclear roles and responsibilities, unclear referral pathways, referral to external networks, limited knowledge of sleep disorders, lack of funding and time constraints, and sceptical views on treatment. Although the majority of the included studies in this review focus on referral barriers, it is equally important to understand what enables the referral. We identified four main categories of referral facilitators to specialist sleep services: clearly defined roles, positive referral experience, utilising external services, and training and education. We then grouped the described barriers and facilitators into five overarching themes: roles and responsibilities, referral pathways, knowledge, funding, and views on treatment.

#### 3.3.1. Roles and responsibilities

Included studies found that poorly defined roles and responsibilities of referring clinicians and sleep specialists hindered timely referral. For example, Hayes et al. (24) reported that generalists’ (family physicians and internists) roles and responsibilities in relation to sleep disorders were unclear, and that they struggled to understand who, when, and where to refer. Screening and referral rather than diagnosis and treatment were seen as an important part of the referring healthcare professionals’ role (24,30). However, sleep specialists were perceived as providing a diagnosis but not treatment which resulted in reluctance to refer patients to sleep specialists (24). This may vary by sleep disorder, although the management of OSA was perceived as a sleep specialist’s responsibility (27), as insomnia care was perceived as within the scope of GPs (29).

Mirroring the identified barriers, clearly defined roles and responsibilities were seen to facilitate referrals to specialist sleep services. A GP’s role was viewed as providing preliminary assessment for at-risk patients and referring to sleep study services, with or without specialist input, for a formal diagnosis (28). Sharing specialty knowledge between nurses and sleep specialists, and training nurses to triage patients was identified as a facilitator to sleep medicine referrals (25).

#### 3.3.2. Referral pathways

Unclear referral pathways were identified as a barrier to referrals to specialist sleep services, whereas a positive experience of the referral process and using external resources were reported to facilitate referrals.

##### 3.3.2.1. Unclear referral pathway

Three studies identified limited understanding of the referral pathways to specialist sleep services as a barrier to referral (28–30), with one study of GPs in Australia also highlighting limited referral options for patients with suspected OSA outside of metropolitan areas (28). Unclear referral and treatment pathways were seen to reduce healthcare professionals’ confidence in discussing sleep issues with patients (30). Although insomnia was perceived as within the scope of primary care, GPs said that they were more likely to refer for co-morbid insomnia with another condition such as anxiety or depression (29). GPs also reported a lack of services and referral pathways for specialist addiction support to assist chronic insomnia patients with benzodiazepine withdrawal (29).

##### 3.3.2.2. Positive referral experience

Easy referrals and good relationships with local sleep services were seen to positively affect the referral process. GPs in a study by Grivell et al. (28) reported that sleep study referrals were straightforward, however, the authors do not describe why this was the case. Similarly, findings from a multi-country study show that some doctors who treat tetraplegia patients appeared satisfied with local sleep services stating good relationships and short waiting times which did not require changing the current referral process (27). It is however unclear from the paper which doctors from which countries the statement refers to. In an US study that explored staff experiences of a new referral initiative within the Department of Veteran Affairs (VA) which serves a different patient population than the Department of Health and Human Services, continuity of sleep specialist care was shown to be improved by limiting reliance on referrals of military veterans, service members, or family members outside the VA healthcare system (25).

##### 3.3.2.3. Utilising external services

There was evidence that using services or providers outside of the referring clinician’s organisation or department might improve access to specialist sleep services. This experience may be country-specific as these issues were reported in studies conducted in Australia and USA and are related to the country’s healthcare funding model. Findings from an Australian study showed that GPs commonly referred to commercial sleep providers to bypass long waiting times, however, the possibility of such referral was dependent on patients’ financial means (28). Additionally, embedding a trained CBTi provider outside the sleep clinics in the USA would have effectively scaled delivery and accelerated access to CBTi/BBTi (Brief Behavioural Therapy for Insomnia) (26). It was recognised that treatment for insomnia in Australia should include best practice psychological treatments such as CBT or mindfulness if accessible through the individual’s healthcare plan (30).

One US study (26) identified factors that acted as a barrier as well as facilitator to referrals to specialist sleep services. The findings show that referral to external networks does not necessarily augment access. In addition, although many providers offered group insomnia treatment which helped to meet the high demand, individual treatment was perceived to achieve better sleep outcomes than group treatment. Using external providers also raised uncertainty about the ethics of private clinics and their potential conflict of interest. GPs were concerned that private providers offer both a diagnosis of OSA and a costly CPAP machine which was identified as a barrier to referral to specialist sleep services (28).

#### 3.3.3. Knowledge

##### 3.3.3.1. Limited knowledge of sleep disorders

Findings from this scoping review show that the referral process is affected by the knowledge of referring healthcare professionals about sleep disorders as well as the perceived limited knowledge of sleep specialists. Sleep health literacy of healthcare professionals was a factor influencing the management and referral to sleep services (30). Many participants could not recall receiving specific training about sleep disorders in their undergraduate or post-graduate courses, but some could remember specific post-graduate training by professional bodies. There seemed to be a disparity in how sleep medicine is viewed. A study of USA’s generalists as well as specialists (24) reported that most generalists did not recognize sleep medicine as a credible sub-specialty and questioned whether specialists know more about sleep disorders than generalists. One of the reasons may be the lack of uniformity in training for sleep specialists. In addition, the perceived lack of value for patients in referring them to specialist sleep centres meant that it was difficult for generalists to convince patients of the referral benefit (24). Another reason why sleep medicine is poorly recognised may be due to the fact that sleep disorders are perceived as a symptom rather than a primary diagnosis by generalists (24,29).

##### 3.3.3.2. Training and education

Raising awareness of sleep issues can positively contribute to the management of sleep disorders and improve access by improving the referral to specialist sleep services. Le Grande et al. (30) raised the importance of improving the education and awareness of sleep issues amongst patients. In addition, improving health professional’s self-efficacy and knowledge regarding sleep issues is also important. There was a belief that additional training and more resources for equipment would allow for the treatment of non-complicated OSA within the unit treating patients with tetraplegia (27). Similarly, targeted education about effective insomnia treatments and referral pathways to specialist services, such as medication withdrawal, would allow for insomnia management within general practice (29). Upskilling other healthcare professionals, such as training nurses to triage, can improve the referral process to the sleep service and can also increase nurses’ autonomy and professional growth (25). In addition, providing educational resources for patients such as videos or pamphlets might improve patient compliance in screening and treatment (30).

#### 3.3.4. Funding

##### 3.1.4.1. Lack of funding and time constraints

Funding issues relating to care for patients with sleep disorders and their potential referral to sleep specialists were identified in Australia (28–30), Europe, UK, Canada, USA, Australia and New Zealand (27). In Australia, there was a recognition that insomnia care can place significant time demands on GPs who are not always financially rewarded for this (29). There was uncertainty about using funded GP Mental Health Treatment Plan for insomnia because insomnia was never considered a primary condition (29). Funding issues were also identified related to Continuous/ Positive Airway Pressure (C/PAP) treatment of OSA in the multi-country study (27). Funding bodies were reported to only accept PAP applications if the patient was diagnosed with a polysomnography and/or by a sleep specialist although these regulations varied between and within the countries included in the multi-country study. Similarly, funding constraints were recognised to affect access to care for OSA since patients on low incomes could not afford CPAP, yet were ineligible for publicly funded CPAP in Australia (28).

Further, time constraints were identified as negatively affecting the referral process. Two studies reported long waiting times to be a barrier to referral to specialist sleep services (27,28). At-risk patients, who may require a referral, can be identified by additional screening for sleep disorders. However, the ability to provide screening was identified as a major barrier by cardiac healthcare professionals due to a lack of time, resources and staff availability. Staff raised concerns about the use of different types of resources e.g. forms and questionnaires which add up to clinicians’ time and can be overwhelming (30).

#### 3.3.5. Views on treatment

The views of the benefit of the treatment may influence clinician’s decision whether to refer a patient to a specialist sleep service or not. A study of physicians involved in the clinical management of OSA in tetraplegia (27) reported conflicting views about the benefits of Positive Airway Pressure (PAP) while routine screening may identify non-symptomatic OSA which was not viewed as requiring treatment (27). Additionally, those doctors who reported referring to sleep specialists for diagnosis and treatment of OSA felt very strongly that OSA should only be managed by a sleep specialist; that it was outside of their scope of practice and that it would be irresponsible to take on management of OSA (27). There was a recognition that it can be challenging for patients to adjust to using CPAP machines and in these instances health professionals can assist with the adjustment period and can inform patients of technology improvements which can make CPAP easier to use (30).

## 4. Discussion

### 4.1. Principal findings

This scoping review identifies and synthesises qualitative evidence relevant to the factors influencing the referral to specialist services for sleep-related disorders. Five overarching themes were identified to influence the referral to specialist sleep services: roles and responsibilities, referral pathways, knowledge, funding, and views on treatment. The identified barriers to referrals to specialist sleep services included unclear roles and responsibilities, unclear referral pathways, referral to external networks, limited knowledge of sleep disorders, lack of funding and time constraints, and sceptical views on treatment. Congruently, facilitators included clearly defined roles, positive referral experience, utilising external services, and training and education. Specific barriers and facilitators varied between countries, healthcare professionals and sleep disorders.

### 4.2. Strengths and limitations of the study

This is the first scoping review offering a comprehensive and systematic search strategy and thorough review of published qualitative evidence on referral barriers and facilitators to specialist sleep services. The main limitation of this study is the small number and heterogeneity in objectives and settings of the included studies. We may not have identified all studies despite our broad inclusion criteria, for example, an additional study was identified during a literature search when writing the discussion section. Further, this scoping review adopted single screening as opposed to the conventional double screening. Given the high yield of the comprehensive searches, using a single reviewer allowed for completing the review in a timely manner. To ensure that the reviewing process was rigorous, a second reviewer was asked to evaluate studies that were not ‘clear cut’ cases. Although a dual review approach is recommended for systematic reviews (31), the approach is resource and labour intensive. When undertaking scoping reviews with limited time resources, single screening is considered to be appropriate (32).

### 4.3. Meaning of the study in the context of existing research

Systematic and scoping reviews of qualitative studies researching healthcare professional perspectives have previously studied barriers and referrals to specialists in the fields of genetic testing (33,34), pulmonary rehabilitation (35), palliative radiotherapy (36), cardiac rehabilitation (37), bariatric surgery (38), lifestyle modification programmes (39), and primary care referrals (40). These systematic reviews identified barriers and facilitators that related to the following themes in our review: Theme 1 Roles and responsibilities (24,25,27–30), Theme 2 Referral pathways (25–30), Theme 3 Knowledge (24,25,27,29,30), Theme 4 Funding (27–30), and Theme 5 Views on treatment (27,30). Barriers and facilitators under these themes were similar to those presented in table 2. For example, one of the identified barriers to referrals to specialist sleep services is the sceptical views of healthcare professionals of the effectiveness and benefits of sleep medicine. Similarly, previous research showed that healthcare professionals’ perception of limited clinical utility and/or effectiveness represented a barrier to referrals for genetic testing (33,34), pulmonary rehabilitation (35), palliative radiotherapy (36), and lifestyle modification programmes (39).

However, scoping reviews in other specialist areas did not identify utilising external services as a facilitator. In addition to the themes we identified, these reviews reported a number of further barriers and facilitators to referrals to specialist services that could be relevant to sleep services. Barriers included poor communication between referrers and specialists (36) and poor communication between clinicians and patients, especially with patients who could not understand English (37). Patient level facilitators included previous experience of referring patients to successful bariatric surgery (38) and patient request to be referred for treatment. Examples of referrer-level facilitators are protected time for providing information such as having time to inform patients about pulmonary rehabilitation and having assistance from other healthcare professionals in identifying suitable patients who would be suitable for referring for pulmonary rehabilitation (35). Several facilitators at the specialist-level have been identified to support the referral. These include interdisciplinary integration and teamwork (39), co-design of programmes with healthcare professionals (39), and ongoing feedback from specialists directly to GPs and other healthcare professionals (39). In addition, marketing of the pulmonary rehabilitation (PR) service such as awareness events, prompts on review template/ computerised pop-ups (making it part of workflow), branded mugs and coasters symbolising PR have been also demonstrated to facilitate referrals for PR (35).

### 4.4. Implications for clinicians and policymakers

In public health and policy, understanding how professional practices change is crucial for implementing effective and evidence-based interventions. It is important to examine conscious as well as subconscious relationships between attitudes and behaviour when proposing policy interventions. Several theoretical models and frameworks exist that aim to understand human behaviour and help identify factors affecting behaviour change, such as the COM-B model and the Theoretical Domains Framework (TDF). The TDF builds on the systems identified in the COM-B model. The Capability Opportunity Motivation-Behaviour (COM-B) model (41) suggests that to perform specific behaviours, we need the knowledge, skills and abilities (capability), the right conditions (opportunity) and the right mindset (motivation). The themes identified in this scoping review align with this model although the included studies investigated them in less depth than the model allows for, since none of the included studies apart from one were theoretically underpinned. The only included study in this scoping review that used a theoretical framework was the multi-country study of OSA management in tetraplegic patients (27) that adopted the TDF framework. For example, while studies in this scoping review clearly identified knowledge (i.e. capability) as a barrier and facilitator for referral, they did not fully investigate the role of opportunity and motivation in determining other barriers and facilitators to referrals.

Nevertheless, findings from this scoping review could be used by clinicians as well as policymakers to make more informed decisions about healthcare planning and delivery for patients with sleep disorders and potentially for other patients requiring specialist care, within these domains. Importantly, there is a need to improve knowledge of sleep disorders and related skills amongst healthcare professionals which can positively impact the referral process. Previous research suggests that there is an urgent need to improve the knowledge of sleep disorders amongst primary care clinicians (8,42,43). Sleep education of healthcare professionals should increase to incorporate the latest research discoveries in sleep medicine which could help raise the profile of sleep medicine as a speciality. Clear referral guidelines with clearly defined roles for healthcare professionals working in different settings would increase capability. Initiatives applied in other settings, such as co-design of referral interventions (39) and positive outreach by specialist centres (35) may increase motivation to refer patients to sleep centres by reducing scepticism about sleep medicine, while minimising unnecessary referrals.

### 4.5. Unanswered questions and future research

The findings of this scoping review provide insight into factors influencing referrals to sleep centres for mainly prevalent sleep disorders (e.g. OSA) in specific settings in a small number of countries. Whilst themes describing barriers and facilitators are shared between countries, settings and sleep disorders, the specific nature of these barriers varies considerably and the existing research lacks an in-depth understanding of the issues. Further research is needed to inform service design in other countries and settings with more in-depth analysis. This review can be used to inform the design of these studies.

## 5. Conclusion

Qualitative research on barriers and facilitators to referrals to specialist sleep services from the referrer’s perspective is limited and heterogenous. Common themes reflect a lack of clarity regarding roles, responsibilities and referral pathways, and limited knowledge of sleep disorders and available treatments. Experiences within these themes vary widely between settings. More robust empirical evidence is therefore needed to help us understand the referrals issues in other countries, settings and related to less common sleep disorders.

## Data Availability

All data produced in the present study are available upon reasonable request to the authors.

## 6. Acknowledgements

The authors would like to thank Jane Falconer, User Support Services Librarian at the London School of Hygiene and Tropical Medicine for her advice on the scoping review protocol.

## 6.2. CRediT authorship contribution statement

**Martina Sykorova**: conceptualization, data curation, formal analysis, investigation, methodology, project administration, visualization, writing – original draft.

**Ellen Nolte**: conceptualization, writing – review and editing.

**Frederick Martineau**: formal analysis, methodology, writing – review and editing.

**Michelle Miller**: supervision, writing – review and editing.

**Sofia Eriksson**: supervision, writing – review and editing

**Ian Smith**: supervision, writing – review and editing.

**Helen Strongman**: conceptualization, data curation, formal analysis, funding acquisition, methodology, supervision, writing – review and editing.

## 6.3. Conflicts of interest

MS has no conflicts of interest to declare.

EN has no conflicts of interest to declare.

FM has no conflicts of interest to declare.

MAM is a voluntary executive member of the British Sleep Society and receives Royalties from two Sleep, Health and Society Textbooks published by Oxford University Press.

SHE has received honoraria for educational activities from Eisai, Fidia, Lincoln and UCB Pharma.

IES has no conflicts of interest to declare.

HS is a voluntary Director and Trustee of Narcolepsy UK, a patient-led charity.

All authors have read and agreed to the published version of the manuscript.

## Appendices

## Appendix A: PRISMA-ScR Checklist

Preferred Reporting Items for Systematic reviews and Meta-Analyses extension for Scoping Reviews (PRISMA-ScR) Checklist

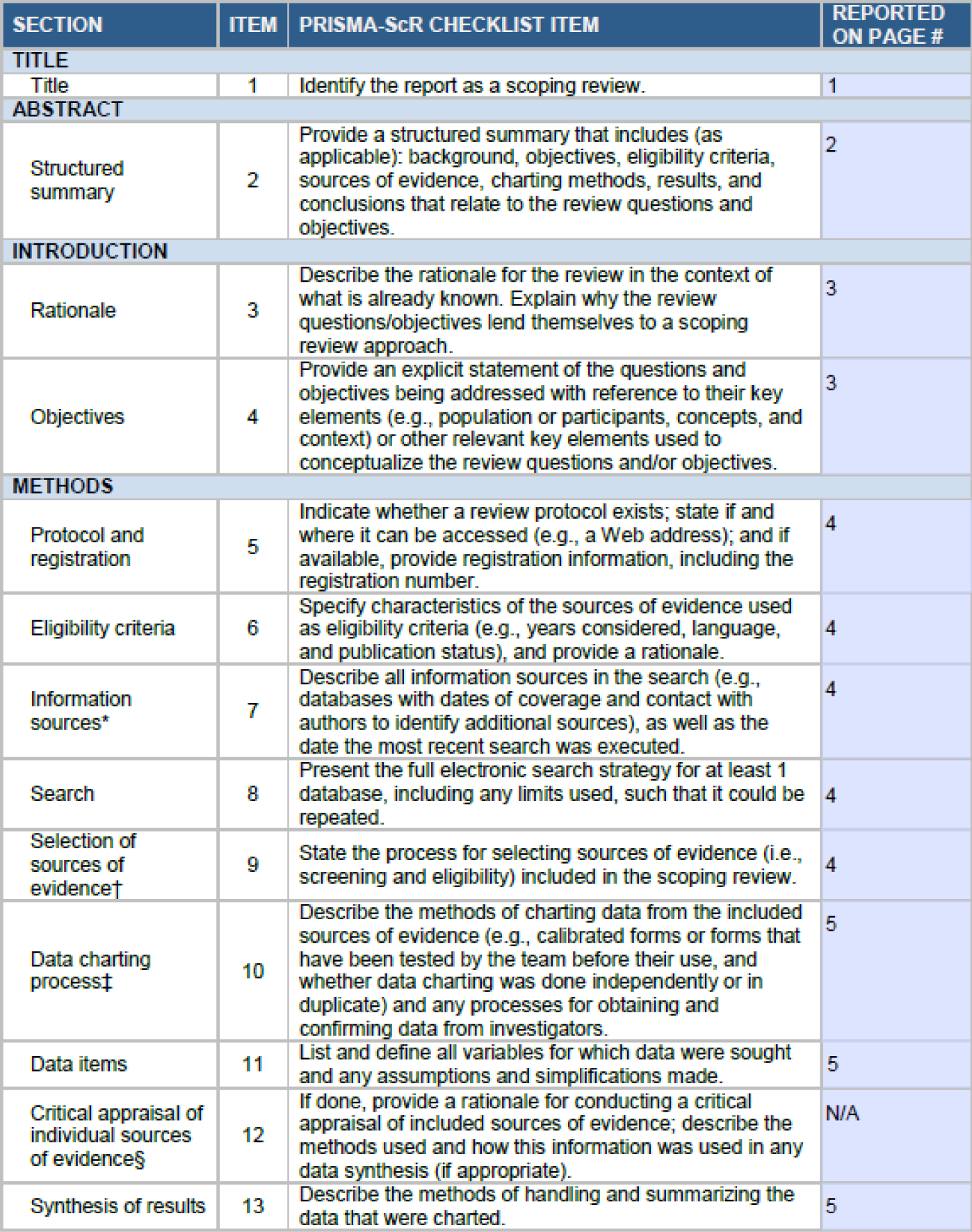
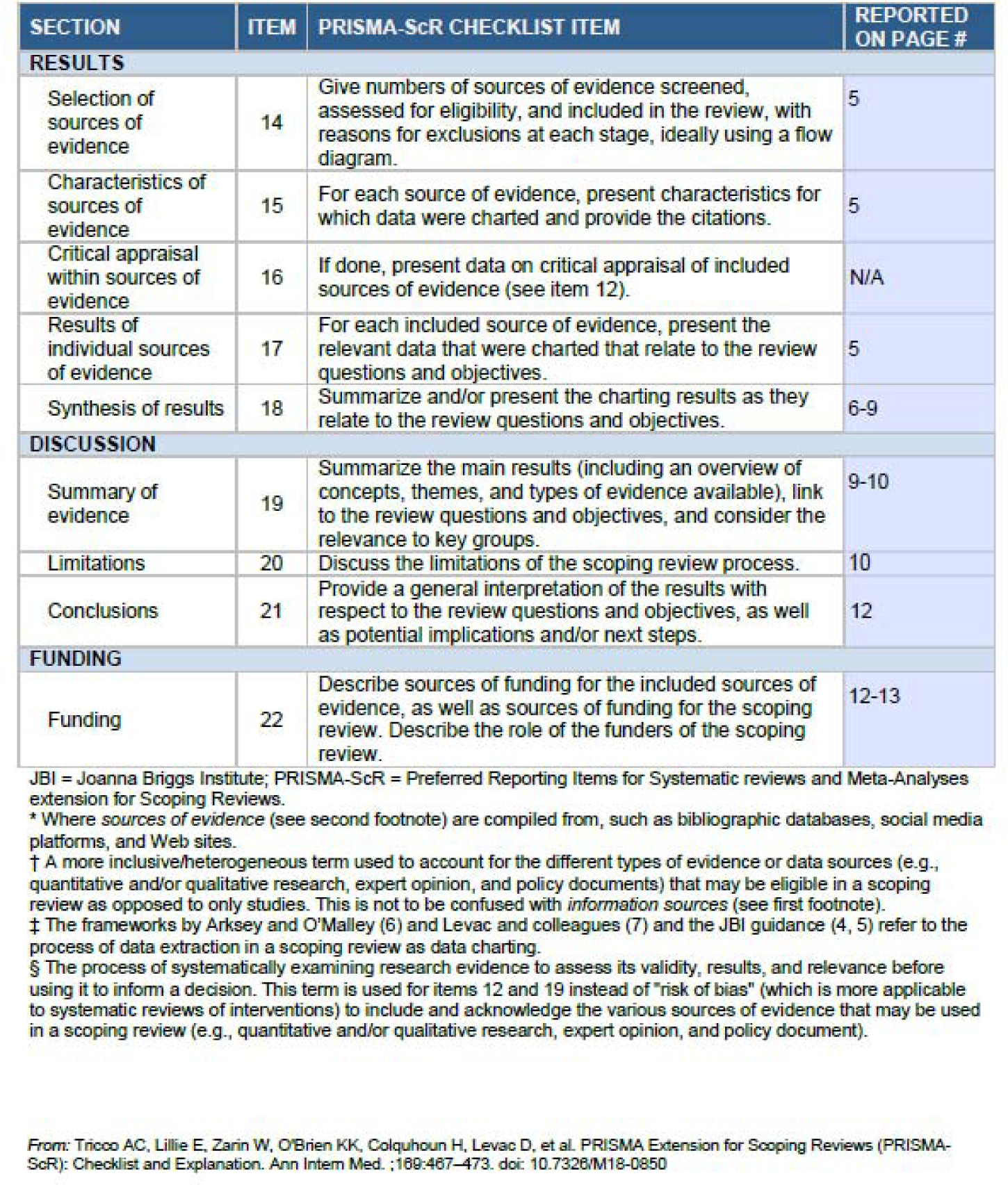

| SECTION | ITEM | PRISMA-ScR CHECKLIST ITEM | REPORTED ON PAGE # |
| --- | --- | --- | --- |
| **TITLE** |  |  |  |
| Title | 1 | Identify the report as a scoping review. | 1 |
| **ABSTRACT** |  |  |  |
| Structured summary | 2 | Provide a structured summary that includes (as applicable): background, objectives, eligibility criteria, sources of evidence, charting methods, results, and conclusions that relate to the review questions and objectives. | 2 |
| **INTRODUCTION** |  |  |  |
| Rationale | 3 | Describe the rationale for the review in the context of what is already known. Explain why the review questions/objectives lend themselves to a scoping review approach. | 3 |
| Objectives | 4 | Provide an explicit statement of the questions and objectives being addressed with reference to their key elements (e.g., population or participants, concepts, and context) or other relevant key elements used to conceptualize the review questions and/or objectives. | 3 |
| **METHODS** |  |  |  |
| Protocol and registration | 5 | Indicate whether a review protocol exists; state if and where it can be accessed (e.g., a Web address); and if available, provide registration information, including the registration number. | 4 |
| Eligibility criteria | 6 | Specify characteristics of the sources of evidence used as eligibility criteria (e.g., years considered, language, and publication status), and provide a rationale. | 4 |
| Information sources* | 7 | Describe all information sources in the search (e.g., databases with dates of coverage and contact with authors to identify additional sources), as well as the date the most recent search was executed. | 4 |
| Search | 8 | Present the full electronic search strategy for at least 1 database, including any limits used, such that it could be repeated. | 4 |
| Selection of sources of evidence† | 9 | State the process for selecting sources of evidence (i.e., screening and eligibility) included in the scoping review. | 4 |
| Data charting process‡ | 10 | Describe the methods of charting data from the included sources of evidence (e.g., calibrated forms or forms that have been tested by the team before their use, and whether data charting was done independently or in duplicate) and any processes for obtaining and confirming data from investigators. | 5 |
| Data items | 11 | List and define all variables for which data were sought and any assumptions and simplifications made. | 5 |
| Critical appraisal of individual sources of evidence§ | 12 | If done, provide a rationale for conducting a critical appraisal of included sources of evidence; describe the methods used and how this information was used in any data synthesis (if appropriate). | N/A |
| Synthesis of results | 13 | Describe the methods of handling and summarizing the data that were charted. | 5 |
| **RESULTS** |  |  |  |
| Selection of sources of evidence | 14 | Give numbers of sources of evidence screened, assessed for eligibility, and included in the review, with reasons for exclusions at each stage, ideally using a flow diagram. | 5 |
| Characteristics of sources of evidence | 15 | For each source of evidence, present characteristics for which data were charted and provide the citations. | 5 |
| Critical appraisal within sources of evidence | 16 | If done, present data on critical appraisal of included sources of evidence (see item 12). | N/A |
| Results of individual sources of evidence | 17 | For each included source of evidence, present the relevant data that were charted that relate to the review questions and objectives. | 5 |
| Synthesis of results | 18 | Summarize and/or present the charting results as they relate to the review questions and objectives. | 6-9 |
| **DISCUSSION** |  |  |  |
| Summary of evidence | 19 | Summarize the main results (including an overview of concepts, themes, and types of evidence available), link to the review questions and objectives, and consider the relevance to key groups. | 9-10 |
| Limitations | 20 | Discuss the limitations of the scoping review process. | 10 |
| Conclusions | 21 | Provide a general interpretation of the results with respect to the review questions and objectives, as well as potential implications and/or next steps. | 12 |
| **FUNDING** |  |  |  |
| Funding | 22 | Describe sources of funding for the included sources of evidence, as well as sources of funding for the scoping review. Describe the role of the funders of the scoping review. | 12-13 |

JB1 = Joanna Briggs Institute; PRISMA-ScR = Preferred Reporting Items for Systematic reviews and Meta-Analyses extension for Scoping Reviews.
\* Where *sources of evidence* (see second footnote) are compiled from, such as bibliographic databases, social media platforms, and Web sites.
† A more inclusive/heterogeneous term used to account for the different types of evidence or data sources (e.g., quantitative and/or qualitative research, expert opinion, and policy documents) that may be eligible in a scoping review as opposed to only studies. This is not to be confused with *information sources* (see first footnote).
‡ The frameworks by Arksey and O'Malley (6) and Levac and colleagues (7) and the JBI guidance (4, 5) refer to the process of data extraction in a scoping review as data charting.
§ The process of systematically examining research evidence to assess its validity, results, and relevance before using it to inform a decision. This term is used for items 12 and 19 instead of "risk of bias" (which is more applicable to systematic reviews of interventions) to include and acknowledge the various sources of evidence that may be used in a scoping review (e.g., quantitative and/or qualitative research, expert opinion, and policy document).
From: Tricco AC, Lillie E, Zarin W, O'Brien KK, Colquhoun H, Levac D, et al. PRISMA Extension for Scoping Reviews (PRISMA-ScR): Checklist and Explanation. *Ann Intern Med.* ;168:467–473. doi: 10.7326/M18-0850

## Appendix B: Scoping review protocol

### Abstract

#### Objective

This scoping review aims to identify and summarise qualitative evidence on barriers and facilitators to referrals to specialist sleep services by healthcare professionals.

#### Introduction

An initial search identified several systematic reviews exploring the enablers and barriers to referrals to a specified service, however, little is known about the qualitative evidence of enablers and barriers to referrals to specialist services for sleep-related disorders from the perspective of healthcare professionals. Unlike quantitative approaches, qualitative approaches can help us understand how, why as well as what factors hinder and facilitate the referral process.

#### Inclusion criteria

The review will consider qualitative studies published in any language between 1995-2024 that explore the barriers and facilitators to referral to specialist services for sleep-related disorders from healthcare care professionals’ perspective. Studies using quantitative methodologies or reporting on the referral issues from patients’ perspectives will be excluded. Animal studies will be also excluded.

#### Methods and analysis

A systematic search of six databases (Cochrane Library, MEDLINE, PubMed, Scopus, EMBASE and Open Grey) will be conducted. A data extraction tool will be used to collect relevant information. The qualitative data will be analysed using a thematic analysis.

#### Discussion

This scoping review summarises factors that influence the referral to specialist sleep services from a healthcare professionals’ perspective. Findings from this study will assist GPs as well as sleep specialists to understand what the current referral issues are and how the referral process could be improved. This study will have implications for clinicians (both generalists and specialists), policy makers, and patients.

#### Ethics and dissemination

Ethics approval is not required. Findings will be disseminated through PPI and professional networks and published in a peer-reviewed scientific journal.

### Introduction

Sleep disorders are a heterogenous group of conditions of diverse aetiology. Although sleep disorders are increasingly recognised as a public health problem worldwide, they remain under-recognised, under-diagnosed and under-treated (The Lancet, 2022). Early diagnosis and treatment by a sleep specialist can reduce many of the negative impacts including poor educational outcomes, unintentional injuries such as road traffic and other accidents as well as wider productivity losses in the workplace (Ramar et al., 2021; The Lancet, 2022).

The gatekeeping system in many countries determines that a patient who requires specialist care needs a referral to sleep medicine services from another healthcare professional (Khatwa et al., 2013; Livingston et al., 2013), which in most cases is a general practitioner (GP) (Saleem et al., 2017; Thorsen et al., 2016; Tzartzas et al., 2019). For example, it is recommended that those who present with symptoms of narcolepsy (Vignatelli et al., 2019) or severe OSA should be referred to a sleep specialist to avoid diagnostic delays (Mansfield et al., 2013). However, there is a variation in the appropriateness and completeness of referrals to specialist sleep services. (Livingston et al., 2013). While it is important to refer those with symptoms of sleep disorders to avoid potential missed cases (Butler and Lee, 2023), inappropriate referrals can result in overwhelming sleep services (Khatwa et al., 2013). A better understanding of the factors that influence referrals to specialist sleep services would therefore improve the diagnosis process and access to appropriate care.

Despite how common sleep disorders are in the general population and the significant impact they have on people and their quality of life, accessing specialist care presents an issue due to limited awareness of these conditions amongst the public as well as healthcare professionals. Access to specialist services, and timely diagnosis and management are crucial in improving the quality of life of people with sleep disorders. However, little is known about the qualitative evidence of enablers and barriers to specialist sleep referrals for people with sleep issues, especially Obstructive Sleep Apnoea (OSA) and narcolepsy.

We conducted a preliminary search of MEDLINE, the Cochrane Database of Systematic Reviews and *JBI Evidence Synthesis* to confirm that there are no current or underway systematic reviews or scoping reviews exploring the factors influencing referrals to specialist sleep services. The objective of this scoping review is to address this research gap by summarising the qualitative evidence exploring barriers and facilitators to referrals to specialist services for sleep-related disorders from healthcare providers’ perspective.

#### Review questions

We used the Participants/Concept/Context framework, as recommended by the Joanna Briggs Institute for scoping reviews (Peters et al., 2022), to structure the primary objective and define the inclusion criteria.

*Participants*: Healthcare professionals referring patients

*Concept*: Qualitative evidence exploring the barriers and facilitators to referrals to specialist services for sleep-related disorders

*Context*: Within the healthcare sector from the perspective of healthcare professionals

##### Research question

1. What is the qualitative evidence exploring the barriers and facilitators to referrals to specialist services for sleep-related disorders from a healthcare providers’ perspective?

#### Inclusion criteria

We will include studies that will meet all of the following criteria:

- Qualitative evidence (\**mixed method studies will be considered*)
- Referrals to specialist sleep/respiratory/neurology/psychiatry services for sleep-related disorders
- Reporting on barriers and/or facilitators to referrals
- Referrals by healthcare professionals/providers
- Referrals for sleep-related disorders
- Studies published between 1995-2024
- Studies in any language

#### Exclusion criteria

We will exclude studies that will meet any of the following criteria:

- Quantitative approaches/evidence
- Reviews, expert opinions, abstracts, editorials, comments, study protocols or no full text available
- Patients’, family members’ or carers’ views/experiences
- Animal studies
- Studies published prior 1995

##### Types of sources

This scoping review will consider all types of qualitative studies data including, but not limited to, designs such as phenomenology, grounded theory, ethnography, qualitative description, interviews, action research, qualitative surveys, case study, participatory research, discourse analysis and narrative research. In addition, systematic reviews that meet the inclusion criteria will also be considered, depending on the research question. Reviews, expert opinions, abstracts, editorials, comments, study protocols or no full text available will not be considered for inclusion in this scoping review.

### Methods

The proposed scoping review will be conducted in accordance with the JBI methodology for scoping reviews (Aromataris et al., 2024) and will be reported using the PRISMA-ScR Checklist (Tricco et al., 2018).

#### Search strategy

The search strategy will aim to locate published studies. A three-step search strategy will be utilized in this review. First an initial limited search of MEDLINE (PubMed) and Google Scholar was undertaken to identify articles on the topic. The text words contained in the titles and abstracts of relevant articles, and the index terms used to describe the articles were used to develop a full search strategy for report the name of the relevant databases/information sources (see appendix 1).

The search strategy, including all identified keywords and index terms, will be adapted for each included database and/or information source. The reference list of all included sources of evidence will be screened for additional studies.

Studies published in any language will be included. Studies published since 1995 when the commercial continuous positive airway pressure (CPAP) machines have entered the market as a treatment option for sleep apnoea. In addition, new discoveries in the treatment of insomnia were made in the 1990s (Neubauer, 2019) and the hypocretin deficiency was established as the cause of narcolepsy in 2000 (Mignot, 2014).

The databases to be searched include Cochrane Library, MEDLINE (via Ovid), PubMed, EMBASE (via Ovid) and Scopus. Sources of grey literature to be searched via the Open Grey database.

#### Study/Source of evidence selection

Following the search, all identified citations will be collated and uploaded into Rayyan (a cloud-based software tool for systematic literature reviews) and duplicates removed. Titles and abstracts will then be screened by one independent reviewer for assessment against the inclusion criteria for the review. Potentially relevant sources will be retrieved in full and their citation details imported into Rayyan. The full text of selected citations will be assessed in detail against the inclusion criteria by one independent reviewer. Reasons for exclusion of sources of evidence at full text that do not meet the inclusion criteria will be recorded and reported in the scoping review. The results of the search and the study inclusion process will be reported in full in the final scoping review and presented in a PRISMA flow diagram (Page et al., 2021).

#### Data extraction

Data will be extracted from papers included in the scoping review by one independent reviewer using a data extraction tool developed by the project team. The data extracted will include specific details about the participants, concept, context, study methods and key findings relevant to the review questions. A draft extraction form is provided (see appendix 2). The draft data extraction tool will be modified and revised as necessary during the process of extracting data from each included evidence source. Modifications will be detailed in the scoping review. Critical appraisal of individual sources of evidence will not be conducted.

### Data analysis and presentation

Data will be analysed thematically and presented in the form of a narrative summary and a table that will describe how the results relate to the reviews objective and research question.

### Author contributions

Author contributions will be reported using the CRediT authorship contribution statement.

## Appendix 1: Search strategy

- Referral
- Service
- Specialist
- Sleep
- Respiratory
- Lung
- Neurology
- Psychiatry
- Sleep disorder
- Sleep disturbance
- Sleep apnoea (US + UK spelling)
- Narcolepsy
- Insomnia
- Fatigue
- Somnipathy
- Parasomnia
- Sleep paralysis
- Sleepwalking
- Hypersomnia
- Dyssomnia
- Restless leg syndrome
- nocturnal myoclonus
- nightmare disorder
- Night terror
- Snoring
- Circadian rhythm disorder
- Nocturia
- Sleepiness
- Excessive Daytime Sleepiness
- Periodic limb movements of sleep
- REM (sleep behaviour) disorder
- Cataplexy
- Tiredness
- Sleep bruxism
- Sleeping beauty syndrome
- Kleine–Levin syndrome
- *familial hibernation syndrome*
- Fatal Familial Insomnia
- *exploding head syndrome*
- *shift work disorder*
- sleep syndrome
- jet lag disorder
- Hypnagogic hallucinations
- Polysomnography
- Respiratory polygraphy
- Qualitative
- Interview
- Observation
- Focus group
- Participation
- Ethnography
- Phenomenology
- Grounded theory
- Case study
- Participatory action
- Interpretative
- Research
- Discourse
- Narrative
- Diary
- Experience
- Belief
- Attitude

## Appendix 2: Draft data extraction tool

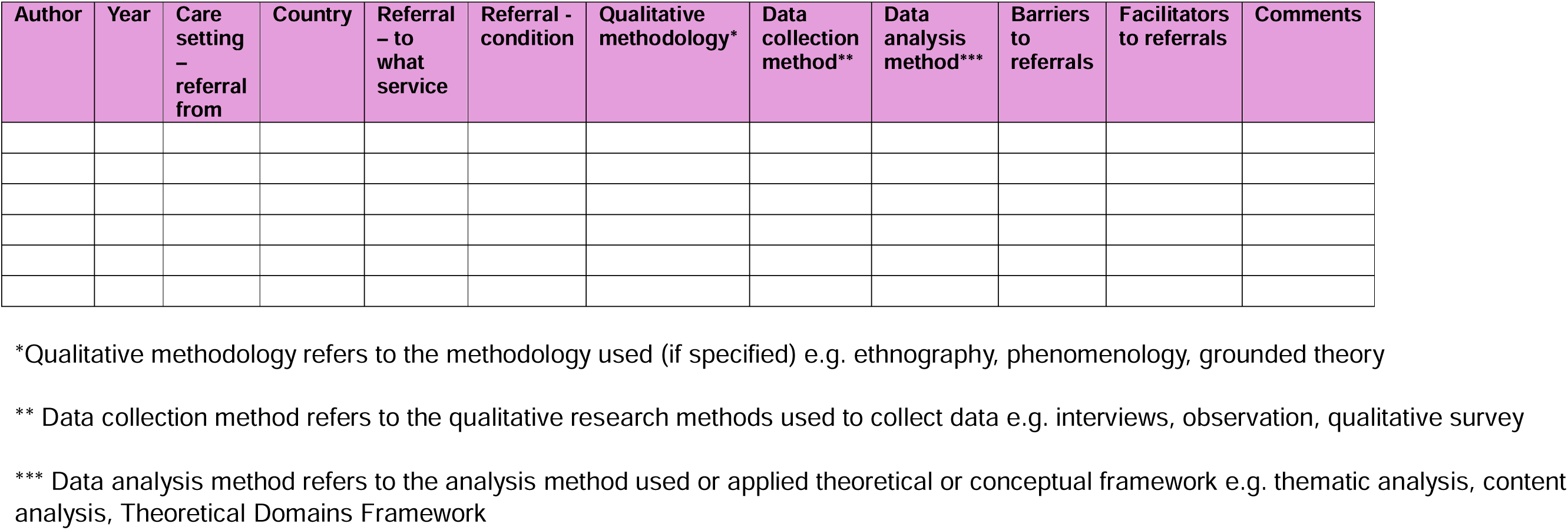

## Appendix C: Search strategy

**Cochrane Library 153**

Search Name: 01.05.2024 v.2

Date Run: 01/05/2024 20:05:18

| ID | Query | Results |
| --- | --- | --- |
| #1 | referral OR access | 59139 |
| #2 | specialist OR service | 61260 |
| #3 | sleep OR respirator* OR lung OR psychiatr* OR neurolog* OR apnea OR apnoea OR narcolepsy OR "sleep disorder" OR "sleep disturbance" OR Insomnia OR "Fatigue" OR "Somnipathy" OR "Parasomnia" OR "Sleep paralysis" OR "Sleepwalking" OR "Hypersomnia" OR "Dyssomnia" OR "Restless leg syndrome" OR "nocturnal myoclonus" OR "nightmare disorder" OR "Night terror" OR "Snoring" OR "Circadian rhythm disorder" OR "Nocturia" OR "Sleepiness" OR "Excessive Daytime Sleepiness" OR "Periodic limb movements of sleep" OR "REM sleep disorder" OR Cataplexy OR tiredness OR "Sleep bruxism" OR "Sleeping beauty syndrome" OR "Kleine Levin syndrome" OR "familial hibernation syndrome" OR "Fatal Familial Insomnia" OR "exploding head syndrome" OR "shift work disorder" OR "sleep syndrome" OR "jet lag disorder" OR "hypnagogic hallucinations" OR Polysomnography OR "Respiratory polygraphy" | 388330 |
| #4 | #1 AND #2 AND 3 | 7982 |
| #5 | qualitative OR Interview OR observ* OR Focus group OR particip* OR ethnograph* OR phenomenolog* OR "grounded theory" OR "case study" OR interpret* OR "action research" OR discourse OR narrative OR Diary OR Experience OR Belief OR Attitude | 807261 |
| #6 | #4 AND #5 | 6878 |
| #7 | referral | 16919 |
| #8 | #7 AND #2 AND #3 | 1367 |
| #9 | #8 AND #5 with Cochrane Library publication date Between Jan 1995 and May 2024 | 1169 |
| #10 | MeSH descriptor: [Referral and Consultation] explode all trees | 3386 |
| #11 | #10 AND #2 AND #3 | 224 |
| #12 | #11 AND #5 | **153** |

Ovid **MEDLINE**(R) ALL <1946 to April 30, 2024> 1672

| ID | Query | Results |
| --- | --- | --- |
| 1 | (referral or access).mp. [mp=title, book title, abstract, original title, name of substance word, subject heading word, floating sub-heading word, keyword heading word, organism supplementary concept word, protocol supplementary concept word, rare disease supplementary concept word, unique identifier, synonyms, population supplementary concept word, anatomy supplementary concept word] | 639107 |
| 2 | (specialist or service or sleep or "sleep specialist" or "sleep service" or "sleep disorder" or "sleep disturbance").mp. [mp=title, book title, abstract, original title, name of substance word, subject heading word, floating sub-heading word, keyword heading word, organism supplementary concept word, protocol supplementary concept word, rare disease supplementary concept word, unique identifier, synonyms, population supplementary concept word, anatomy supplementary concept word] | 788125 |
| 3 | (respirator* or psychiatr* or neurolog*).mp. [mp=title, book title, abstract, original title, name of substance word, subject heading word, floating sub-heading word, keyword heading word, organism supplementary concept word, protocol supplementary concept word, rare disease supplementary concept word, unique identifier, synonyms, population supplementary concept word, anatomy supplementary concept word] | 1630155 |
| 4 | (sleep or respirator* or psychiatr* or neurolog*).mp. [mp=title, book title, abstract, original title, name of substance word, subject heading word, floating sub-heading word, keyword heading word, organism supplementary concept word, protocol supplementary concept word, rare disease supplementary concept word, unique identifier, synonyms, population supplementary concept word, anatomy supplementary concept word] | 1849219 |
| 5 | (specialist or service).mp. [mp=title, book title, abstract, original title, name of substance word, subject heading word, floating sub-heading word, keyword heading word, organism supplementary concept word, protocol supplementary concept word, rare disease supplementary concept word, unique identifier, synonyms, population supplementary concept word, anatomy supplementary concept word] | 535015 |
| 6 | 1 and 4 and 5 | 8037 |
| 7 | (referral or access or investigation).mp. [mp=title, book title, abstract, original title, name of substance word, subject heading word, floating sub-heading word, keyword heading word, organism supplementary concept word, protocol supplementary concept word, rare disease supplementary concept word, unique identifier, synonyms, population supplementary concept word, anatomy supplementary concept word] | 1320385 |
| 8 | 4 and 5 and 7 | 9146 |
| 9 | (sleep or lung or respirator* or psychiatr* or neurolog*).mp. [mp=title, book title, abstract, original title, name of substance word, subject heading word, floating sub-heading word, keyword heading word, organism supplementary concept word, protocol supplementary concept word, rare disease supplementary concept word, unique identifier, synonyms, population supplementary concept word, anatomy supplementary concept word] | 2663602 |
| 10 | 5 and 7 and 9 | 9918 |
| 11 | (referral or access or investigation).mp. [mp=title, book title, abstract, original title, name of substance word, subject heading word, floating sub-heading word, keyword heading word, organism supplementary concept word, protocol supplementary concept word, rare disease supplementary concept word, unique identifier, synonyms, population supplementary concept word, anatomy supplementary concept word] | 1320385 |
| 12 | (specialist or service).mp. [mp=title, book title, abstract, original title, name of substance word, subject heading word, floating sub-heading word, keyword heading word, organism supplementary concept word, protocol supplementary concept word, rare disease supplementary concept word, unique identifier, synonyms, population supplementary concept word, anatomy supplementary concept word] | 535015 |
| 13 | (sleep or respirator* or lung or psychiatr* or neurolog*).mp. [mp=title, book title, abstract, original title, name of substance word, subject heading word, floating sub-heading word, keyword heading word, organism supplementary concept word, protocol supplementary concept word, rare disease supplementary concept word, unique identifier, synonyms, population supplementary concept word, anatomy supplementary concept word] | 2663602 |
| 14 | 11 and 12 and 13 | 9918 |
| 15 | (qualitative or Interview or observ* OR Focus group or particip* or ethnograph* or phenomenolog* or "grounded theory" or "case study" or interpret* or "action research" or discourse or narrative or Diary or Experience or Belief or Attitude).mp. [mp=title, book title, abstract, original title, name of substance word, subject heading word, floating sub-heading word, keyword heading word, organism supplementary concept word, protocol supplementary concept word, rare disease supplementary concept word, unique identifier, synonyms, population supplementary concept word, anatomy supplementary concept word] | 3812635 |
| 16 | 14 and 15 | 3652 |
| 17 | (sleep or respirator* or lung or psychiatr* or neurolog* or apnea or apnoea or narcolepsy or "sleep disorder" or "sleep disturbance" or Insomnia or "Fatigue" or "Somnipathy" or "Parasomnia" or "Sleep paralysis" or "Sleepwalking" or "Hypersomnia" or "Dyssomnia" or "Restless leg syndrome" or "nocturnal myoclonus" or "nightmare disorder" or "Night terror" or "Snoring" or "Circadian rhythm disorder" or "Nocturia" or "Sleepiness" or "Excessive Daytime Sleepiness" or "Periodic limb movements of sleep" or "REM sleep disorder" or Cataplexy or tiredness or "Sleep bruxism" or "Sleeping beauty syndrome" or "Kleine Levin syndrome" or "familial hibernation syndrome" or "Fatal Familial Insomnia" or "exploding head syndrome" or "shift work disorder" or "sleep syndrome" or "jet lag disorder" or "hypnagogic hallucinations" or Polysomnography or "Respiratory polygraphy").mp. [mp=title, book title, abstract, original title, name of substance word, subject heading word, floating sub-heading word, keyword heading word, organism supplementary concept word, protocol supplementary concept word, rare disease supplementary concept word, unique identifier, synonyms, population supplementary concept word, anatomy supplementary concept word] | 2801504 |
| 18 | 11 and 12 and 17 | 10315 |
| 19 | 15 and 18 | 3845 |
| 20 | (referral or access or investigation).mp. [mp=title, book title, abstract, original title, name of substance word, subject heading word, floating sub-heading word, keyword heading word, organism supplementary concept word, protocol supplementary concept word, rare disease supplementary concept word, unique identifier, synonyms, population supplementary concept word, anatomy supplementary concept word] | 1320385 |
| 21 | limit 20 to (abstracts and yr="1995 -Current") | 1126528 |
| 22 | exp *Referral and Consultation/ | 35397 |
| 23 | limit 22 to (abstracts and yr="1995 -Current") | 20326 |
| 24 | *Referral and Consultation/ | 28334 |
| 25 | limit 24 to (abstracts and yr="1995 -Current") | 15397 |
| 26 | referral.mp. or "Referral and Consultation"/ | 186187 |
| 27 | limit 26 to (abstracts and yr="1995 -Current") | 146155 |
| 28 | 12 and 13 and 26 | 5264 |
| 29 | 12 and 17 and 26 | 5404 |
| 30 | 15 and 29 | **1672** |

**PubMed**

(referral[Title/Abstract]) OR (consultation and referral[MeSH Terms])

PubMed **(referral[Title/Abstract]) OR (consultation and referral[MeSH Terms])**
| ID | Query | Results |
| --- | --- | --- |
| #1 | Search: (referral[Title/Abstract]) OR (consultation and referral[MeSH Terms]) | 195,701 |
| #2 | Search: specialist OR service | 2,090,871 |
| #3 | Search: sleep OR respirator* OR lung OR psychiatr* OR neurolog* OR apnea OR apnoea OR narcolepsy OR "sleep disorder" OR "sleep disturbance" OR Insomnia OR "Fatigue" OR "Somnipathy" OR "Parasomnia" OR "Sleep paralysis" OR "Sleepwalking" OR "Hypersomnia" OR "Dyssomnia" OR "Restless leg syndrome" OR "nocturnal myoclonus" OR "nightmare disorder" OR "Night terror" OR "Snoring" OR "Circadian rhythm disorder" OR "Nocturia" OR "Sleepiness" OR "Excessive Daytime Sleepiness" OR "Periodic limb movements of sleep" OR "REM sleep disorder" OR Cataplexy OR tiredness OR "Sleep bruxism" OR "Sleeping beauty syndrome" OR "Kleine Levin syndrome" OR "familial hibernation syndrome" OR "Fatal Familial Insomnia" OR "exploding head syndrome" OR "shift work disorder" OR "sleep syndrome" OR "jet lag disorder" OR "hypnagogic hallucinations" OR Polysomnography OR "Respiratory polygraphy" | 4,093,863 |
| #4 | Search: ((#1) AND (#2)) AND (#3) | 15,136 |
| #5 | Search: qualitative OR Interview OR observ* OR Focus group OR particip* OR ethnograph* OR phenomenolog* OR "grounded theory" OR "case study" OR interpret* OR "action research" OR discourse OR narrative OR Diary OR Experience OR Belief OR Attitude | 9,016,356 |
| #6 | Search: (#4) AND (#5) | 7,441 |
| #7 | Search: referral[MeSH Major Topic] | 35,397 |
| #8 | Search: referral[Title/Abstract] | 132,437 |
| #9 | Search: ((#7) AND (#2)) AND (#3) | 3,523 |
| #10 | Search: (#9) AND (#5) | 1,624 |
| #11 | Search: ((#8) AND (#2)) AND (#3) | 9,100 |
| #12 | Search: (#11) AND (#5) | 4,850 |
| #13 | Search: (#9) AND (#5) Filters: **from 1995 - 2024** | 1,332 |

**EMBASE**

*\*excluded Medline results*

Embase Classic+Embase <1947 to 2024 April 30>

| ID | Query | Results |
| --- | --- | --- |
| 1 | referral.mp. or exp patient referral/ | 304855 |
| 2 | limit 1 to (abstracts and "remove medline records" and yr="1995 - 2024") | 117449 |
| 3 | ("sleep service" or "sleep specialist" or "respirator* service" or "respirator* specialist" or "lung service" or "lung specialist" or "psychiatr* service" or "psychiatr* specialist" or "neurolog* service" or "neurolog* specialist").mp. [mp=title, abstract, heading word, drug trade name, original title, device manufacturer, drug manufacturer, device trade name, keyword heading word, floating subheading word, candidate term word] | 5870 |
| 4 | limit 3 to (abstracts and "remove medline records" and yr="1995 - 2024") | 2798 |
| 5 | (apnea or apnoea or narcolepsy or "sleepADJdisorder" or "sleepADJdisturbance" or Insomnia or "Fatigue" or "Somnipathy" or "Parasomnia" or "SleepADJparalysis" or "Sleepwalking" or "Hypersomnia" or "Dyssomnia" or "RestlessADJlegADJsyndrome" or "nocturnalADJmyoclonus" or "nightmareADJdisorder" or "NightADJterror" or "Snoring" or "CircadianADJrhythm" or "Nocturia" or "Sleepiness" or "ExcessiveADJDaytimeADJSleepiness" or "PeriodicADJlimbADJmovementsADJofADJsleep" or "REMADJsleepADJdisorder" or Cataplexy or tiredness or "SleepADJbruxism" or "SleepingADJbeautyADJsyndrome" or "KleineADJLevinADJsyndrome" or "familialADJhibernationADJsyndrome" or "FatalADJFamilialADJInsomnia" or "explodingADJheadADJsyndrome" or "shiftADJworkADJdisorder" or "sleepADJsyndrome" or "jetADJlagADJdisorder" or "hypnagogicADJhallucinations" or Polysomnography or "RespiratoryADJpolygraphy").mp. [mp=title, abstract, heading word, drug trade name, original title, device manufacturer, drug manufacturer, device trade name, keyword heading word, floating subheading word, candidate term word] | 630372 |
| 6 | limit 5 to (abstracts and "remove medline records" and yr="1995 - 2024") | 232668 |
| 7 | 2 and 4 and 6 | 94 |
| 8 | 2 and 4 | 406 |
| 9 | (qualitative or Interview or observ* or Focus group or particip* or ethnograph* or phenomenolog* or "grounded theory" or "case study" or interpret* or "action research" or discourse or narrative or Diary or | 10435646 |
|  | Experience or Belief or Attitude).mp. [mp=title, abstract, heading word, drug trade name, original title, device manufacturer, drug manufacturer, device trade name, keyword heading word, floating subheading word, candidate term word] |  |
| 10 | limit 9 to (abstracts and "remove medline records" and yr="1995 - 2024") | 3286263 |
| 11 | 8 and 10 | **132** |

**SCOPUS**

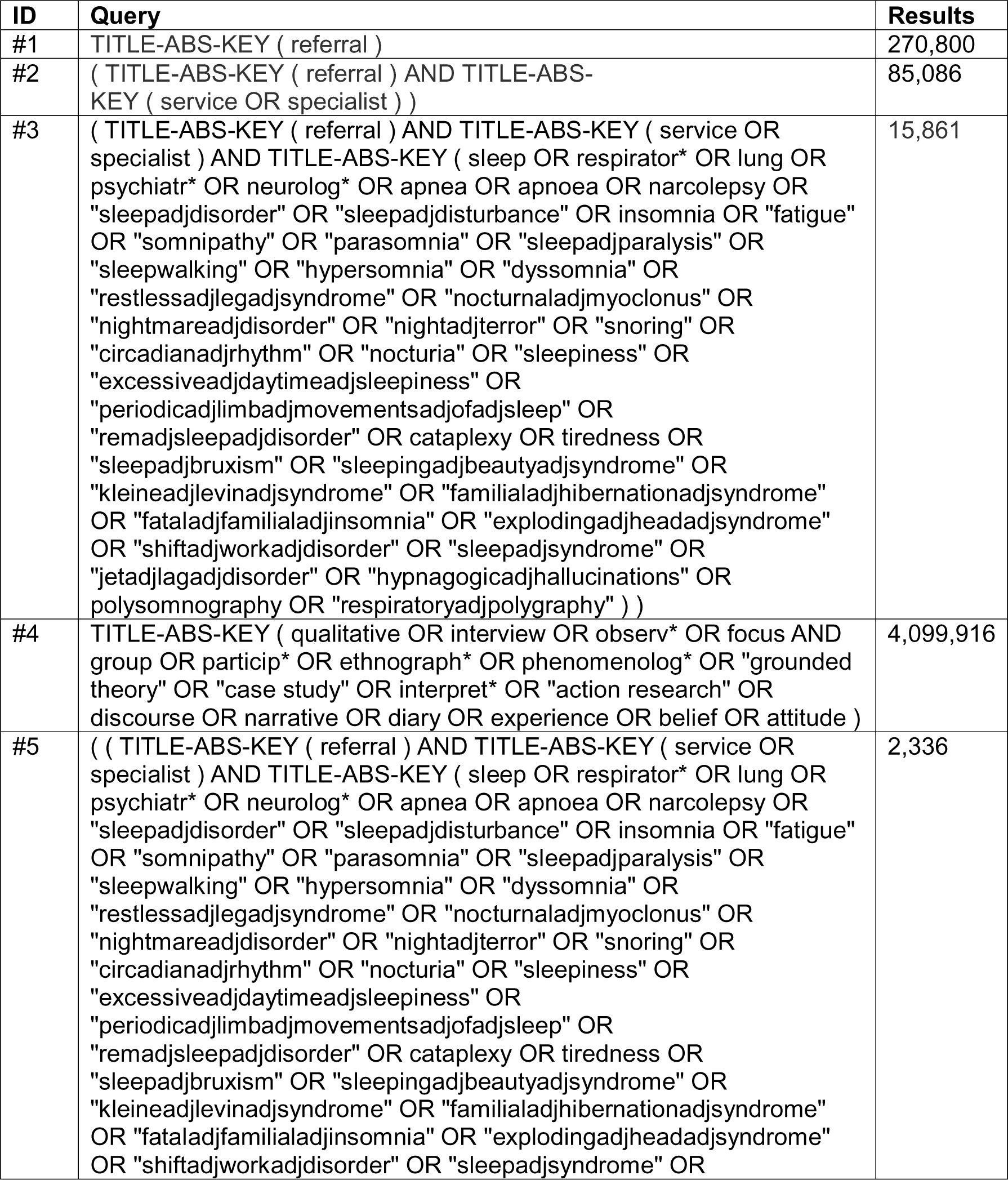
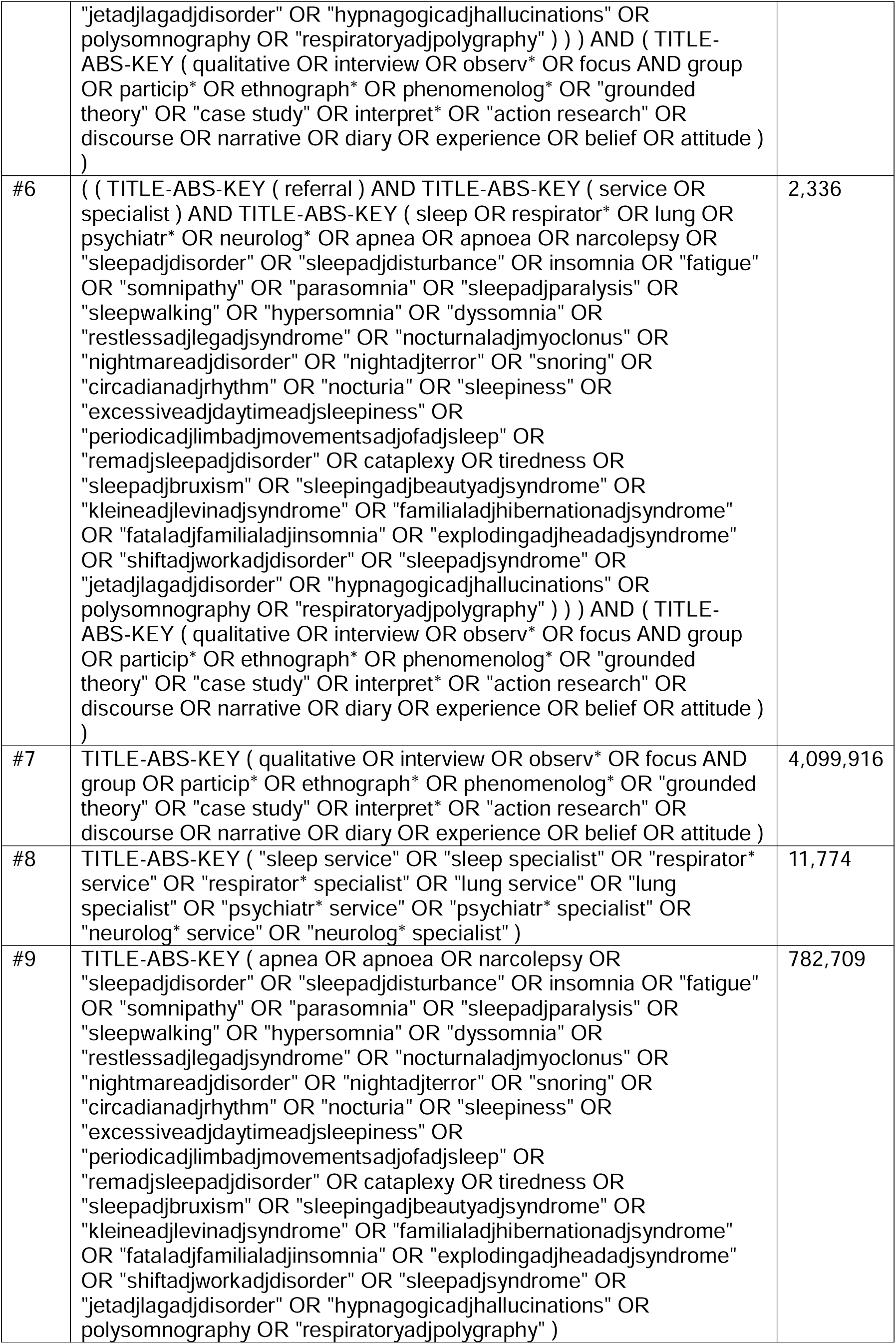
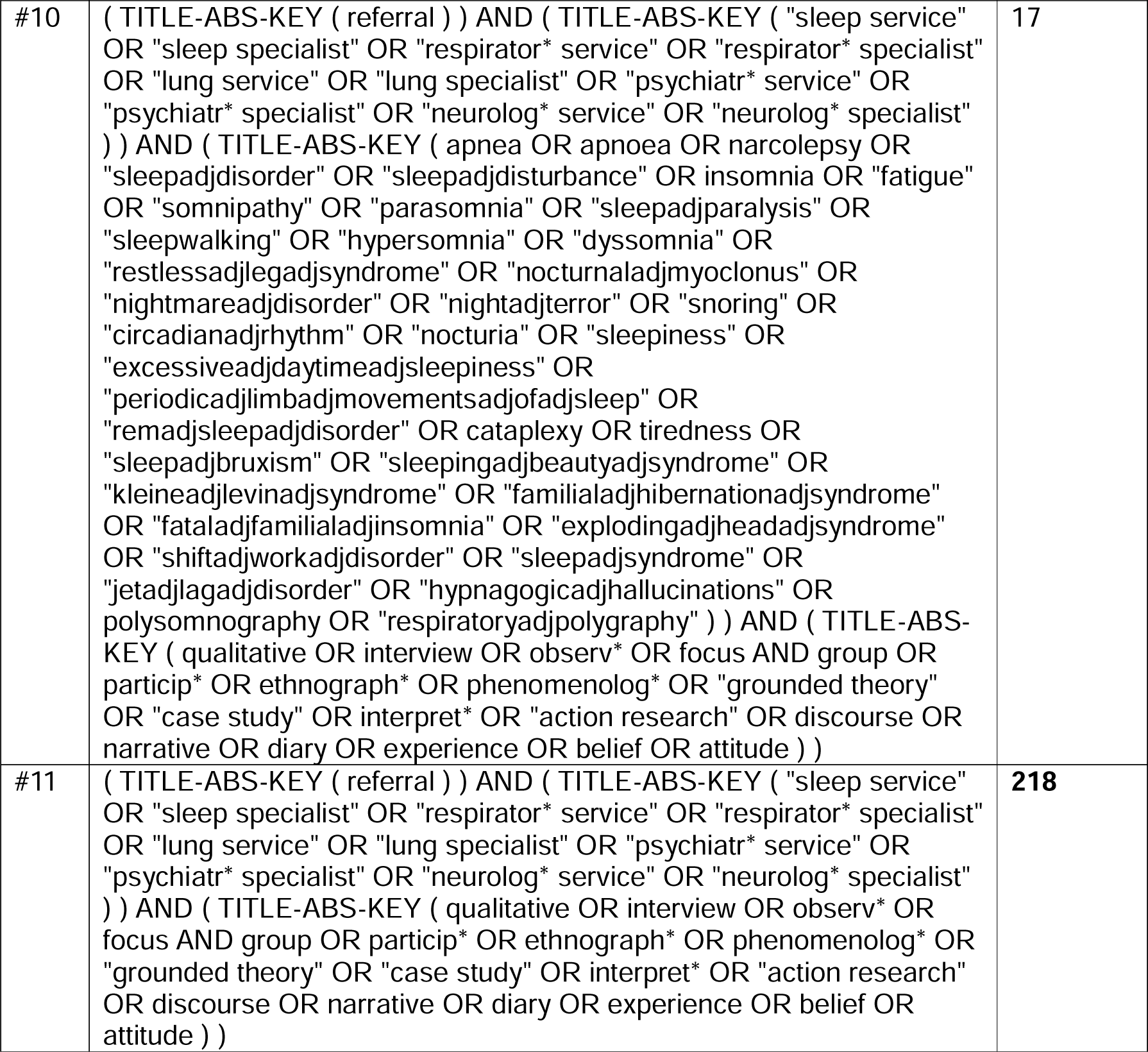

**Open Grey database is no longer being added to after 2020.**

0 documents found

## Appendix D: Data extraction tool

|  |  |
| --- | --- |
| Rayyan no. | = Rayyan ID number |
| Title | = title of the article |
| **Author/s** | = *list of authors* |
| **Year** | = *year published* |
| **Country** | = *where was study conducted* |
| **Referral from** | = *referring clinician/s* |
| **Referral - condition** | = *type of sleep disorder/s* |
| **Sample size** | = *number of participants* |
| **Qualitative methodology** | = *refers to the methodology used (if specified) e.g. ethnography, phenomenology, grounded theory* |
| **Data collection method** | = *refers to the qualitative research methods used to collect data e.g. interviews, observation, qualitative survey* |
| **Data analysis method** | = *refers to the analysis method used or applied theoretical or conceptual framework e.g. thematic analysis, content analysis, TDF* |
| **Barriers to referrals** | = *identified barriers* |
| **Facilitators to referrals** | = *identified facilitators* |
| **Facilitator as well as barrier** | = *identified barriers & facilitators* |
| **Comments** | = *any additional information/ comments* |

